# Normal Variability of Urinary Markers in Individuals with Spinal Cord Injury and Disease Using Intermittent Catheterization

**DOI:** 10.64898/2026.02.02.26345397

**Authors:** Rochelle E. Tractenberg, Suzanne L. Groah, Ethan Newcomb, Christopher Riegner, Catherine Forster

## Abstract

Accurate diagnosis of urinary tract infection (UTI) in individuals with neurogenic lower urinary tract dysfunction (NLUTD) due to spinal cord injury or disease (SCI/D) remains a major clinical challenge. Standard diagnostic tools—including urine dipstick, urinalysis, and culture—lack population-specific reference ranges, and existing criteria are often insufficient to distinguish infection from background variability. To address this gap, we conducted a longitudinal study to define the range of “normal” variability in common urinary biomarkers among individuals with SCI/D who manage their bladders using intermittent catheterization (IC). Participants with NLUTD due to SCI/D who manage their bladders with IC provided urine samples at least two weeks apart, while asymptomatic. We assessed urinary white blood cell count, nitrite, leukocyte esterase, culture-based findings, and urine neutrophil gelatinase-associated lipocalin (uNGAL) to characterize intra-individual stability and inter-individual variation in biomarker profiles. Findings demonstrate that urine parameters exhibit measurable but bounded variability in the absence of UTI, and that deviations beyond these thresholds may support more accurate and individualized UTI diagnosis. By defining the normal range within which values vary without the emergence of symptoms, we hope to further inform clinical and researcher decision-making around variability that moves an individual beyond their normal range of variation in these urinary markers. Operationalizing biologic variability can reduce diagnostic uncertainty and improve antimicrobial stewardship in this frequently overtreated population.

## Introduction

Urinary tract infection (UTI) is the most common outpatient infection worldwide, and for people with spinal cord injury or disease (SCI/D) and neurogenic lower urinary tract dysfunction (NLUTD), it is the most common infection, secondary condition, cause for emergency room visits, and infectious cause of hospitalization^1–3^. Recent research has demonstrated that “gold standard” diagnostic tests, specifically urinalysis (UA) and standard urine culture (SUC), have limited utility for identifying UTI in this population^4,5^. Table 1 (reprinted with permission^6^) outlines variability across several guidelines and recommendations relating to the identification of UTI in individuals with NLUTD due to SCI/D (with shading indicating inconsistencies).

**Table 1.** Variability in definitions for diagnosis of UTI in SCI (adapted from Tractenberg et al. 2025^6^).

|  | **Authoritative Guideline** |  | **Diagnostic Approach Suggestions** |  |  |
| --- | --- | --- | --- | --- | --- |
|  | **NIDRR**<br>**Consensus**<br>**Statement**<br>**(1992)<sup>7</sup>** | **IDSA (2009)<sup>8</sup>**<br>**& EAU**<br>**(2024)<sup>9</sup>** | **SCI-High UTI Project**<br>**(2019)<sup>10</sup>** |  | **Pediatric UTI in**<br>**SB (2013)<sup>11</sup>** |
|  |  |  | Inpatient | Outpatient |  |
| Definition /<br>Criteria | - Symptoms<br>- Inflammation<br>- Bacteria | - Symptoms<br>- Bacteria | - Bladder<br>symptoms<br>+ Fever<br>- Bacteria | - Bladder<br>symptoms<br>- + general<br>symptoms<br>- Bacteria | - Symptoms<br>- Inflammation<br>- Bacteria |
| Symptoms | >1 symptom: | >1 symptom: |  |  | number of<br>symptoms<br>unspecified |
|  |  | Fever | Fever (variable<br>definitions) |  | Fever |
|  | ↑ Spasticity | ↑ Spasticity |  | ↑ Spasticity |  |
|  | Autonomic<br>dysreflexia | Autonomic<br>dysreflexia |  | AD (not<br>standard<br>definition) |  |
|  |  | Rigors |  |  |  |
|  | Dysuria | Dysuria |  |  | Dysuria |
|  | ↑ Incontinence |  |  | ↑<br>Incontinence | ↑ Incontinence |
|  |  | Urgent<br>urination |  |  |  |
|  |  | Acute<br>hematuria | Unprovoked new gross<br>hematuria |  |  |
|  | Smelly <u>and</u><br>cloudy urine |  | Change in urine odor<br><u>and</u> clarity |  | Smelly <u>and/or</u><br>cloudy urine |
|  |  |  |  | Vomiting |  |
|  | Malaise | Malaise |  | Malaise |  |
|  | Sense of unease | Sense of unease |  |  |  |
|  | Lethargy | Lethargy |  |  |  |
|  |  | Mental status change |  |  |  |
|  | Pain over kidney | Flank pain |  |  | New back pain |
|  | Bladder pain | Suprapubic pain |  |  |  |
|  |  | Pelvic pain |  |  |  |
|  |  |  |  |  | Abdominal pain |
|  |  |  | Orthostasis (outpatients) |  |  |
| Pyuria | Leukocytes in urine |  |  |  |  |
|  | Pyuria (>10/HPF) |  |  | New pyuria (amount undefined) | Pyuria (>10/HPF) |
| Bacteria | 100 CFU/mL if IC; 10,000 CFU/mL if condom catheter; Any if indwelling catheter | >=1,000 CFU/mL | Not directly addressed, recommend:<br>>100 CFU/mL if IC<br>>10,000 CFU/mL if void<br>Any detectable if suprapubic aspirate |  | >100,000 CFU/mL of a single organism |
| Notes: Table adapted from Tractenberg et al. 2025<sup>6</sup>. Grey shading identifies discordance.<br>AD: Autonomic dysreflexia; HPF: high powered field; CFU = colony forming units; mL: milliliter; IC: intermittent catheter |  |  |  |  |  |

In addition to the incongruence amongst guidances, standard diagnostic assessments (dipstick, UA, SUC) have low sensitivity and specificity for UTI in this population. For example, individuals with SCI/D may have chronic urothelial inflammation, reflected in greater than expected levels of white blood cells (WBC) in the urine,^12,13^ in the absence of symptoms. Nitrites in urine are indicative of the presence of certain Gram-negative organisms, many of which are present to a greater extent in the urine of people with SCI/D^14^. Finally, people with SCI/D have a high prevalence of asymptomatic bacteriuria^15,16^. The guidance documents shown in Table 1 each note that, while the recommendations are based on substantial clinical experience of the participating panelists, they lack high-quality evidence to support the recommendations. Each document notes the need for research that can generate evidence to support decision making around UTI diagnosis.

To advance evidence and clinical care around UTI among adults with NLUTD due to SCI/D, we sought to describe the natural variation in standard of care UTI assessments as well as an emerging potential biomarker for UTI. The objective of this project is to describe the range of urinary markers for UTI specific to the SCI/D population using intermittent catheterization (IC) to manage their bladders, in the absence of symptoms of UTI, so that stronger evidence about biomarkers for UTI can be identified.

## Methods

### Study Design and Patient Population

This is prospective cohort study of “normal variability” in urinary markers among adults with NLUTD due to SCI/D. Based on observed variability over time and across people in these markers, we sought to explore reasonable thresholds for “change from normal” that might inform future validation of individualized UTI diagnosis. We recruited people aged 18 and older with SCI/D due to either SCI, multiple sclerosis or spina bifida who use IC for bladder management, live in the community (i.e. not in a long-term care facility), who had a history of 2 or more episodes of cloudy or smelly urine in the prior year at time of consent, and who lived in the Metropolitan Washington, DC area between August 2020-October 2024. Our exclusion criteria included: 1) use of oral or intravesical prophylactic antibiotics; 2) psychologic or psychiatric conditions influencing the ability to follow instructions; 3) use of oral or intravenous antibiotics within the past 2 weeks; 4) sexual activity within the previous 72 hours; 5) menstruating at the time of collection; and 6) participation in another study with which results could be confounded. We designed the study to obtain two urine samples, separated by at least two weeks, from individuals who were asymptomatic for 72 hours at the time of sampling. We defined “asymptomatic” as the absence of any symptoms on the Urinary Symptom Questionnaire for Neuropathic Bladder-Intermittent Catheter version^17^ (USQNB-IC; described below) for 72 hours *before and after* the time of urine sampling. IRB approval was obtained from our institution (#1124) and informed consent was obtained from all participants prior to inclusion in the study.

### Symptom Assessment

We used the USQNB-IC, a validated tool^17^ designed for use by patients, researchers, and clinicians, to assess symptoms in this study. Participants completed the USQNB-IC at least 4 times: at the time of each urine collection (reporting on symptoms over the prior 72 hours), and once on each of the three days after each urine collection to ensure that urine samples reflect durability of at least 72 hours of an asymptomatic state, and that participants are “truly asymptomatic” and not “asymptomatic but about to become symptomatic”. Endorsement of any USQNB-IC symptom within the 72 hours prior to the sample collection resulted in rescheduling the sample collection for a later time. If the participant endorsed any of the USQNB-IC symptoms during the 72 hours after the urine acquisition, the urine already collected was considered “pre-symptomatic”, and a repeat urine sampling was scheduled following the same method.

### Sample Collection and Processing

We collected a 50-100 ml urine sample from a new, unused intermittent catheter under sterile conditions. After collection, urine samples were stored at 4 degrees Celsius until processing, which was within 8 hours of collection. The urine was then aliquoted for use in multiple assays.

14 ml of urine was used for UA and SUC. UA, urine microscopy, and SUC was completed used standard laboratory techniques. 9 ml of urine was aliquoted for uNGAL measurement. The urine for uNGAL analysis was centrifuged and the supernatant was aliquoted and subsequently frozen at -80 degrees Celsius within 8 hours of initial collection. There were no freeze-thaw cycles between the time when the samples were initially frozen and when uNGAL was measured. All samples were thawed at the same time for batch measurement. uNGAL was measured by ELISA (Bioporto, Grusbakken, Denmark) in duplicate at the Forster Lab at University of Pittsburgh.

### Urine Sample Characterization

To define normal “variability” within person, we first characterized urine samples in terms of their symptoms over 72 hours, urine inflammatory markers, and cultivable bacteria according to five, non-mututally exclusive groups, shown in Table 2. Groups 1 and 2 are defined based on symptoms plus UA/SUC result, whereas groups 3-5 are based solely on how long the participant remained asymptomatic. Participants in group 3 were asymptomatic on two consecutive collections, whereas group 4 was asymptomatic for single, non-consecutive, collections. Participants in group 5 were asymptomatic at the time of sample collection but then developed at least one symptom within 72 hours. Of note, the groups are not mutually exclusive, and a sample can be placed in more than one group. All participants were asymptomatic at the time the sample was collected; all groups but one (“presymptomatic”) reflect asymptomatic state for 72 hours after the sample.

**Table 2.** Classification of Urine Samples Based on (lack of) Symptoms, UA, and SUC in Individuals with NLUTD due to SCI/D.

| Group | Label | Definition | Notes |
| --- | --- | --- | --- |
| Group 1 | **Standards-Negative Asymptomatic<sup>†</sup> (S)** | Individual is asymptomatic for 72h, UA reveals no pyuria (WBC 0–5), LE negative or trace, nitrite negative, any SUC growth *below* 10<sup>5</sup> CFU. | Most stringent and conservative definition of “healthy urine”, featuring symptoms, UA and SUC results. |
| Group 2 | **UA-Negative Asymptomatic<sup>†</sup> (N)** | Individual is asymptomatic for 72h, UA without pyuria (WBC 0–5), LE negative or trace, and nitrite negative. SUC not considered. | Aligns with clinical thresholds for urine lacking indicators of “inflammation”. |
| Group 3<sup>‡</sup> | **Stable Asymptomatic<sup>†</sup> (C)** | Two samples (at least 2 weeks apart) from the same individual who remains asymptomatic for 72 hours on both samples; UA/SUC not considered. | An individual who is stably asymptomatic. |
| Group 4<sup>‡</sup> | **Single Asymptomatic<sup>†</sup> (A)** | Individual is asymptomatic for 72h after single sample. UA/SUC not considered. | Threshold for “asymptomatic” designation. |
| Group 5<sup>‡</sup> | **Pre-symptomatic<sup>†\*</sup> (P)** | Individual is asymptomatic at time of sample but develops symptoms within 72h. UA/SUC not considered. | Reflects latent or subclinical activity potentially leading to UTI; could be early indicator of microbial or inflammatory shift. |

Notes.
UA= urinalysis; SUC = standard urine culture. UTI = urinary tract infection. WBC = white blood cell count. LE = leukocyte esterase. CFU = colony forming units. 72h = for 72 hours after sample.
<sup>†</sup>“Urinary symptoms” defined as any of the 29 symptoms on the Urinary Symptoms Questionnaire for Neurogenic Bladder- Intermittent Catheter user (USQNB-IC) version.
<sup>‡</sup> Depends on symptoms only. Given the equivocality about the use/utility of urinalysis and standard urine culture for the diagnosis of a UTI<sup>18</sup>, definitions of “healthy” urine that feature results from these methods are not necessarily realistic.
\* No participant gave a urine sample if they endorsed any symptoms when the sample was scheduled to happen. Sample collection was rescheduled in this case.

Data collection utilized REDCap. Data analyses focused on descriptive statistics only, utilizing SPSS v. 29.0 (2024, IBM)^19^; RStudio version 2025.05.1+513^20^ was used for visualizations.

## Results

Our goal was to obtain two samples at least two weeks apart from individuals who remained asymptomatic for 72 hours before and after sample collection. In 10 cases, a participant did remain asymptomatic after sample collection, but either UA (n=4) or SUC (n=12) were not available, requiring a third (or 4th) sample to be collected. Of these, only three had unknown (n=1) or incomplete (n=2) classifications. One sample was UA negative and had no SUC available, so could have met criteria for both Standards Negative and UA Negative (S+N),but was characterized as UA negative (N) only. Two samples were missing UA; one had <10^5^ growth, so might have met S+N, and the other had nothing reported in the SUC output (but SUC was done), so neither S nor N could be determined.

In one case, the participant began the study, but the study ended before they could complete their second sample. All samples that met inclusion criteria (listed above) were included. Thus, number of samples per person ranges from 1-4.

Demographics: Table 3 presents the descriptive statistics for all 99 participants who met inclusion and exclusion criteria and from whom at least one urine sample was obtained. Demographics are presented for all participants together and then stratified according to whether at least one of their samples (or two in the “stable asymptomatic” classification) met each classification of the urine based on symptoms, UA, and SUC. Individuals may appear in multiple strata because their samples met the classification criteria.

**Table 3.**
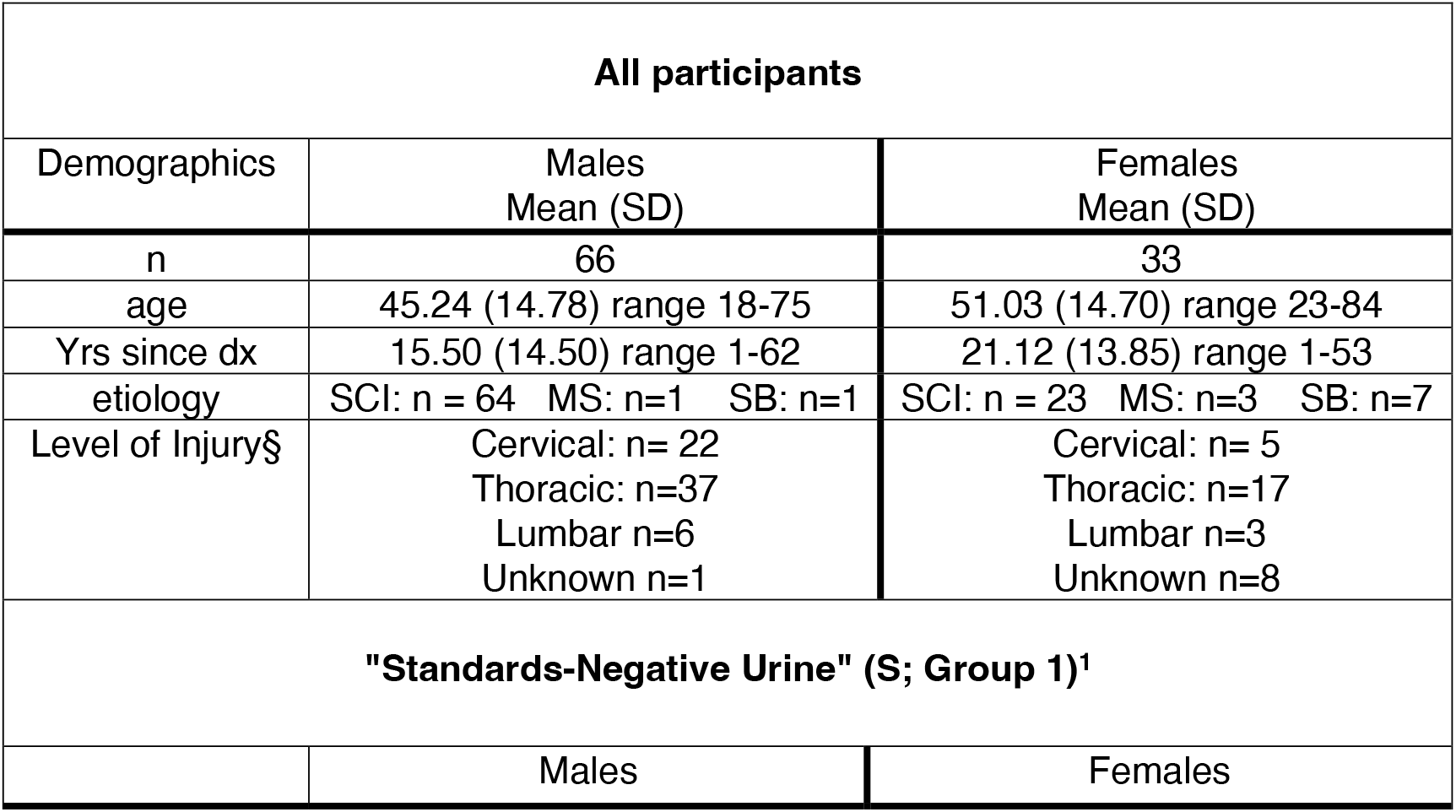
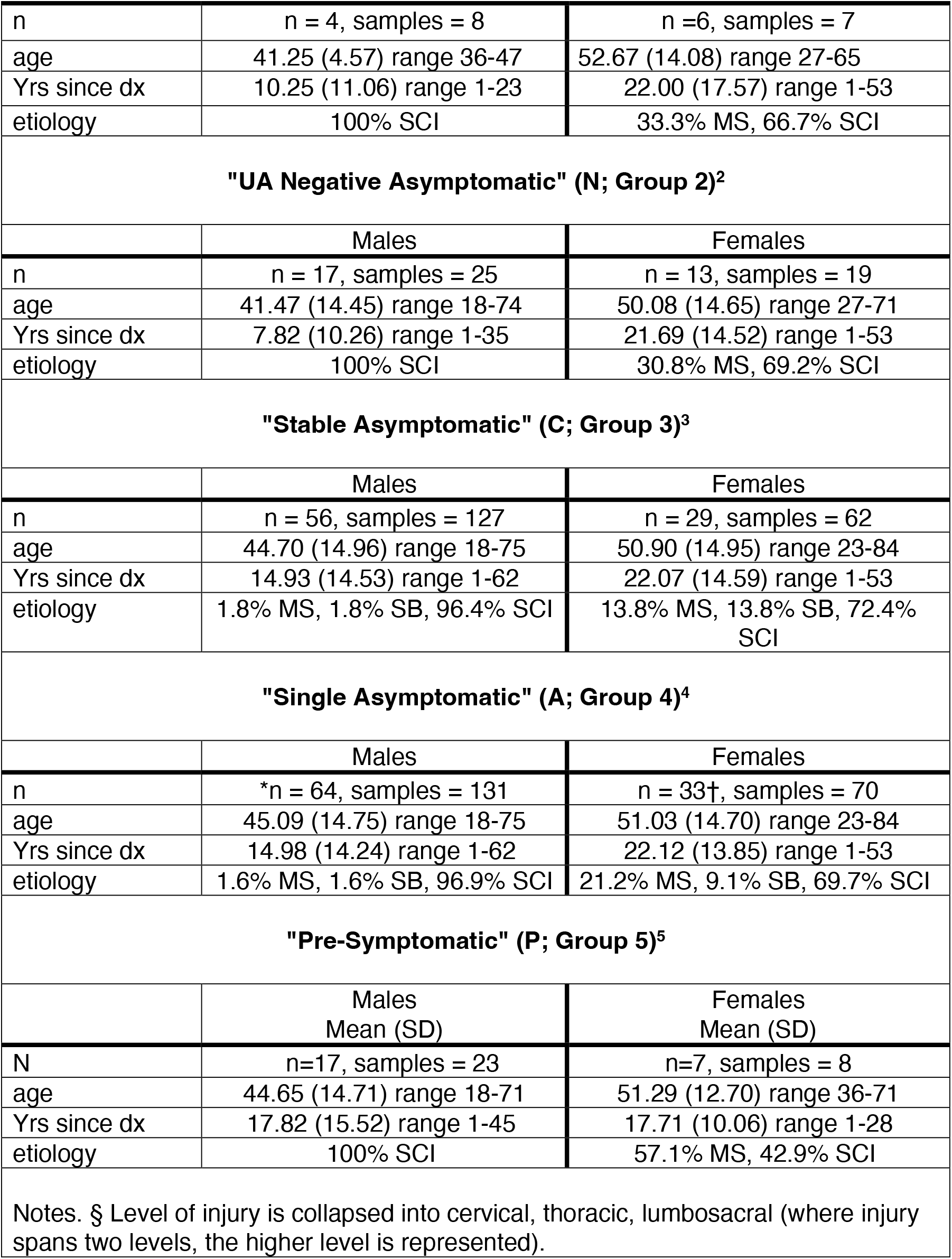
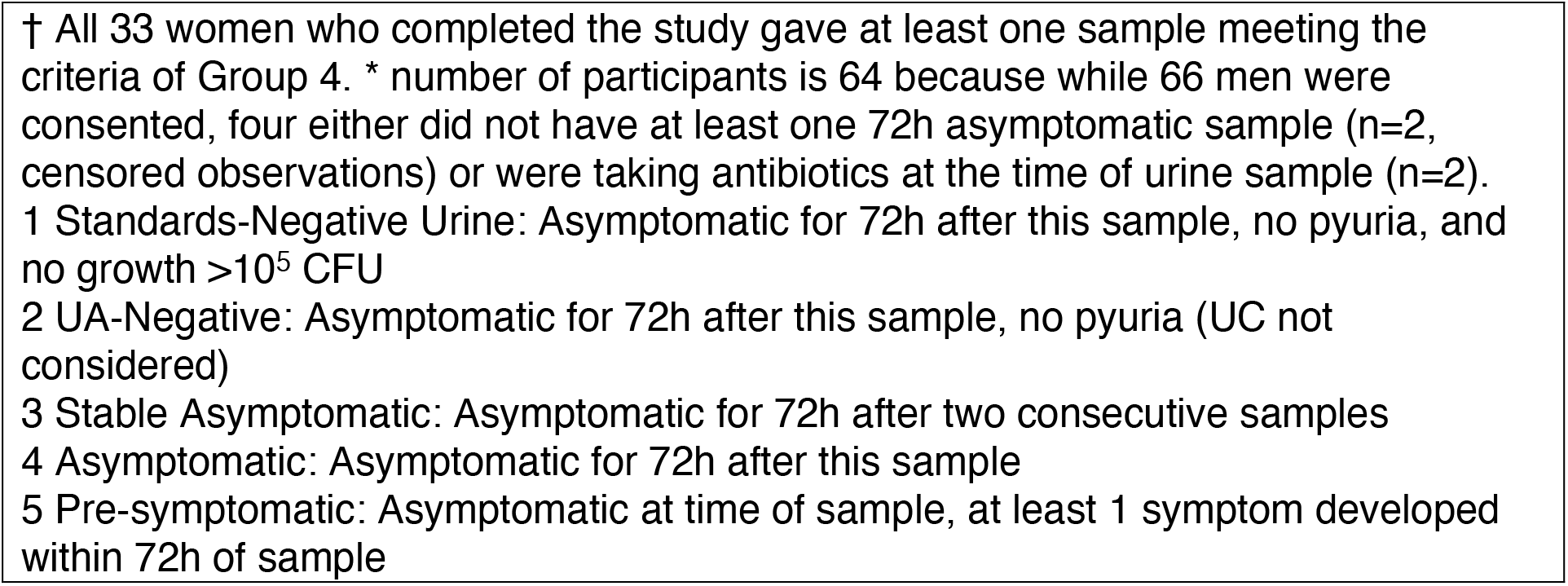
Demographic information for all participants, and for participants with urine samples falling into each of the five Groups.

| All participants |  |  |
| --- | --- | --- |
| Demographics | Males<br>Mean (SD) | Females<br>Mean (SD) |
| n | 66 | 33 |
| age | 45.24 (14.78) range 18-75 | 51.03 (14.70) range 23-84 |
| Yrs since dx | 15.50 (14.50) range 1-62 | 21.12 (13.85) range 1-53 |
| etiology | SCI: n = 64 MS: n=1 SB: n=1 | SCI: n = 23 MS: n=3 SB: n=7 |
| Level of Injury§ | Cervical: n= 22<br>Thoracic: n=37<br>Lumbar n=6<br>Unknown n=1 | Cervical: n= 5<br>Thoracic: n=17<br>Lumbar n=3<br>Unknown n=8 |
| "Standards-Negative Urine" (S; Group 1)<sup>1</sup> |  |  |
|  | Males | Females |
| n | n = 4, samples = 8 | n =6, samples = 7 |
| age | 41.25 (4.57) range 36-47 | 52.67 (14.08) range 27-65 |
| Yrs since dx | 10.25 (11.06) range 1-23 | 22.00 (17.57) range 1-53 |
| etiology | 100% SCI | 33.3% MS, 66.7% SCI |
| **"UA Negative Asymptomatic" (N; Group 2)<sup>2</sup>** |  |  |
|  | Males | Females |
| n | n = 17, samples = 25 | n = 13, samples = 19 |
| age | 41.47 (14.45) range 18-74 | 50.08 (14.65) range 27-71 |
| Yrs since dx | 7.82 (10.26) range 1-35 | 21.69 (14.52) range 1-53 |
| etiology | 100% SCI | 30.8% MS, 69.2% SCI |
| **"Stable Asymptomatic" (C; Group 3)<sup>3</sup>** |  |  |
|  | Males | Females |
| n | n = 56, samples = 127 | n = 29, samples = 62 |
| age | 44.70 (14.96) range 18-75 | 50.90 (14.95) range 23-84 |
| Yrs since dx | 14.93 (14.53) range 1-62 | 22.07 (14.59) range 1-53 |
| etiology | 1.8% MS, 1.8% SB, 96.4% SCI | 13.8% MS, 13.8% SB, 72.4% SCI |
| **"Single Asymptomatic" (A; Group 4)<sup>4</sup>** |  |  |
|  | Males | Females |
| n | *n = 64, samples = 131 | n = 33†, samples = 70 |
| age | 45.09 (14.75) range 18-75 | 51.03 (14.70) range 23-84 |
| Yrs since dx | 14.98 (14.24) range 1-62 | 22.12 (13.85) range 1-53 |
| etiology | 1.6% MS, 1.6% SB, 96.9% SCI | 21.2% MS, 9.1% SB, 69.7% SCI |
| **"Pre-Symptomatic" (P; Group 5)<sup>5</sup>** |  |  |
|  | Males<br>Mean (SD) | Females<br>Mean (SD) |
| N | n=17, samples = 23 | n=7, samples = 8 |
| age | 44.65 (14.71) range 18-71 | 51.29 (12.70) range 36-71 |
| Yrs since dx | 17.82 (15.52) range 1-45 | 17.71 (10.06) range 1-28 |
| etiology | 100% SCI | 57.1% MS, 42.9% SCI |
| Notes. § Level of injury is collapsed into cervical, thoracic, lumbosacral (where injury spans two levels, the higher level is represented). |  |  |

† All 33 women who completed the study gave at least one sample meeting the criteria of Group 4. \* number of participants is 64 because while 66 men were consented, four either did not have at least one 72h asymptomatic sample (n=2, censored observations) or were taking antibiotics at the time of urine sample (n=2).
1 Standards-Negative Urine: Asymptomatic for 72h after this sample, no pyuria, and no growth $>10^5$ CFU
2 UA-Negative: Asymptomatic for 72h after this sample, no pyuria (UC not considered)
3 Stable Asymptomatic: Asymptomatic for 72h after two consecutive samples
4 Asymptomatic: Asymptomatic for 72h after this sample
5 Pre-symptomatic: Asymptomatic at time of sample, at least 1 symptom developed within 72h of sample

Figure 1 shows the proportions of urine samples contributed by female and male participants that fall into each Group of classification.

**Figure 1.**
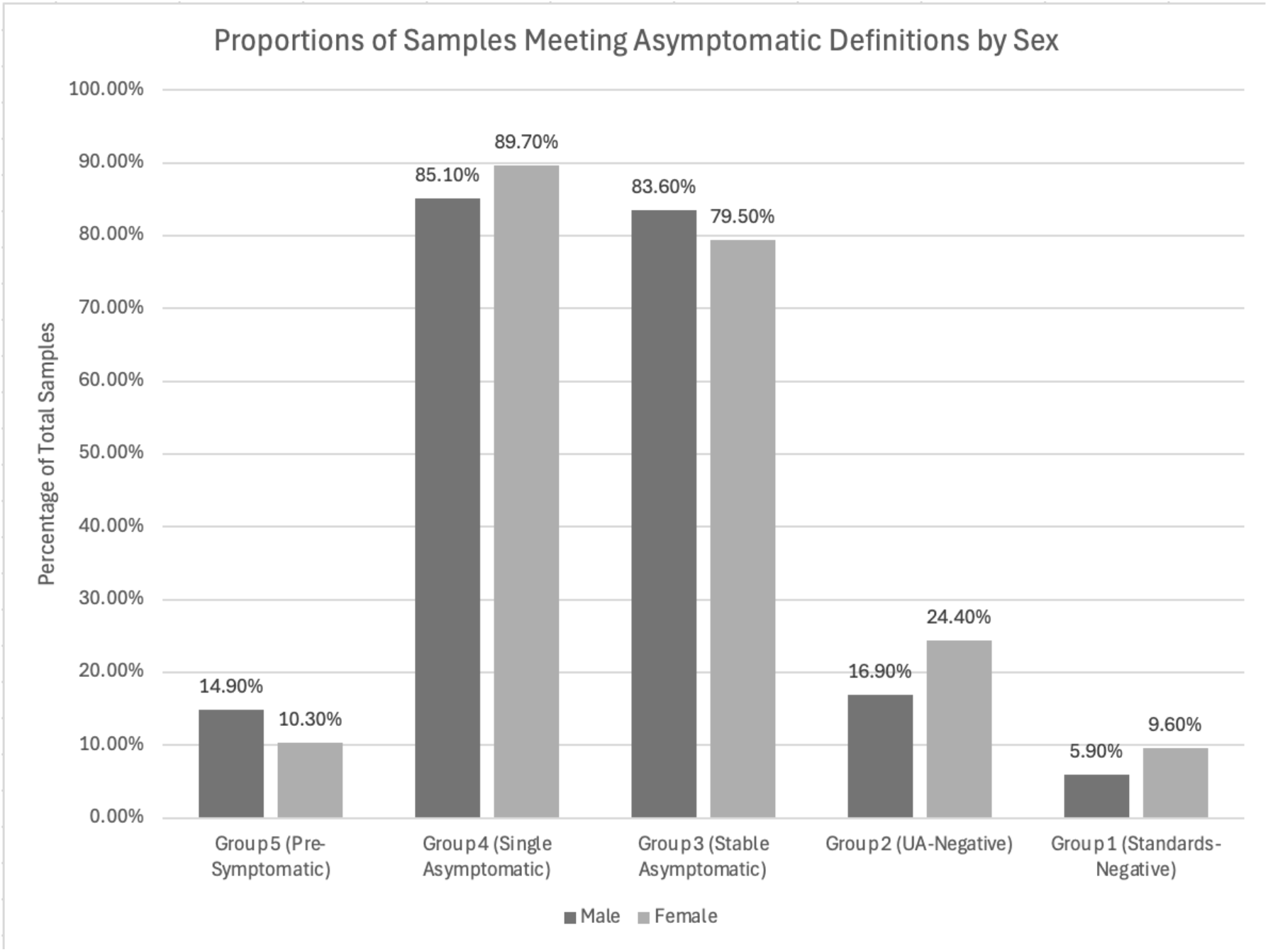
Proportions of urine samples meeting each Group, by sex.

Marginally more samples met the less strict (one time) than more strict (two successive times) asymptomatic criteria. Neither individuals nor samples were compared inferentially because samples could fall into more than one category, and individuals cannot be categorized because many participants contributed samples falling into more than one category.

For each of these categories, we calculated the percentages of individual samples meeting the aforementioned definitions for each of the common urinary markers (or minimum, maximum, and 95th percentile values). These are presented in Table 4.

**Table 4.**
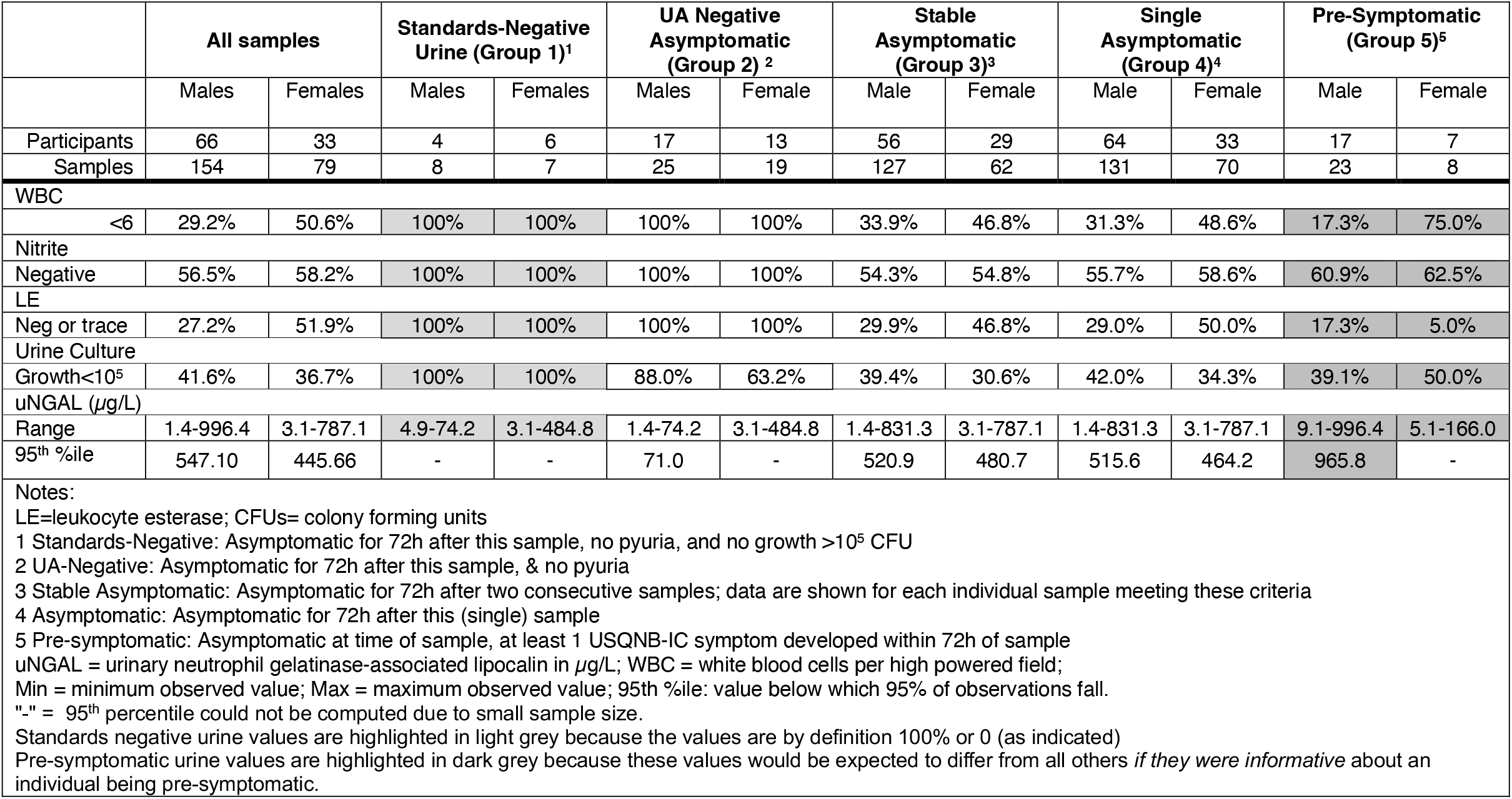
Distributions for urinary markers by sex, stratified by classification based on symptoms, urinalysis results, and standard urine culture results.

In Table 4, Standards negative (group 1) urine values are highlighted in light grey because the values are by definition 100% or 0 (as indicated), while Pre-symptomatic (group 5) urine values are highlighted in dark grey because these values would be expected to differ from all others if they were informative about an individual being pre-symptomatic.

In the asymptomatic state, the majority of samples from males and females with NLUTD due to SCI/D have SUC growth >10^5^ cfu/mL, have nitrite detectable on UA, and have urine WBC ≥6 cells/hpf. In contrast, of all the asymptomatic samples, only 6.4% (15/233 samples) were UA negative as well as having SUC growth <10^5^ cfu/mL, and only 12.9% (30/233) were UA negative without considering SUC results. This clearly indicates limited utility of “positive” results (per this study defined as urine WBC >=6/hpf, nitrite presence, bacterial growth >10^5^ cfu/mL) of standard urine assessment (UA/SUC) among this population.

Across all samples, males tended to have greater pyuria and LE compared to females. This difference in markers of inflammation was most notable in participants when their samples were Pre-Symptomatic, as 78% of samples from males had pyuria compared to only 25% of samples from females. There was little difference in nitrite positivity based on sex, with 54-62% of samples being negative for nitrites (note that Groups 1 and 2 by definition have negative UAs). In terms of bacteriuria, 60% of samples from males met our criteria for bacteriuria, which was stable regardless of symptoms (Cultures were positive in 58.0% of single asymptomatic samples, 60.6% of two consecutive asymptomatic samples, and 60.0% of presymptomatic samples). By contrast, samples from females had a slightly higher rate of bacteriuria: Cultures were positive in 65.7% of single asymptomatic samples, 69.4% of samples that was one of two consecutive samples, and 50% of the presymptomatic samples. Finally, although central tendencies for uNGAL were not reported due to significant skewing of the data, the male samples in the Pre-symptomatic Group (Group 5) had the highest 95^th^ percentile uNGAL value compared to all other groups. Samples that were UA Negative (Group 2) and Standards-Negative (Group 1) in males had the lowest peak uNGAL value (74.2) of any group, although this was a very small group of samples. However, while Presymptomatic samples from males had their highest top value (996.4), the top value of Presymptomatic samples from females was their *lower* (166.0) compared with other categories.

Samples were reclassified into four mutually-exclusive groups to better understand within-individual variability across multiple samples. We eliminated the classification for asymptomatic on successive samples (Group 2) and classified every sample according to the ‘lowest’ group for which they met all criteria. For example, no sample that was both UA negative and Standards negative was included in the UA negative-only group.

Figure 2 shows the variability across participants’ 2-4 samples in terms of the lowest group definition (Level 1 being negative on both UA and Standards) their samples met over time.

**Figure 2.**
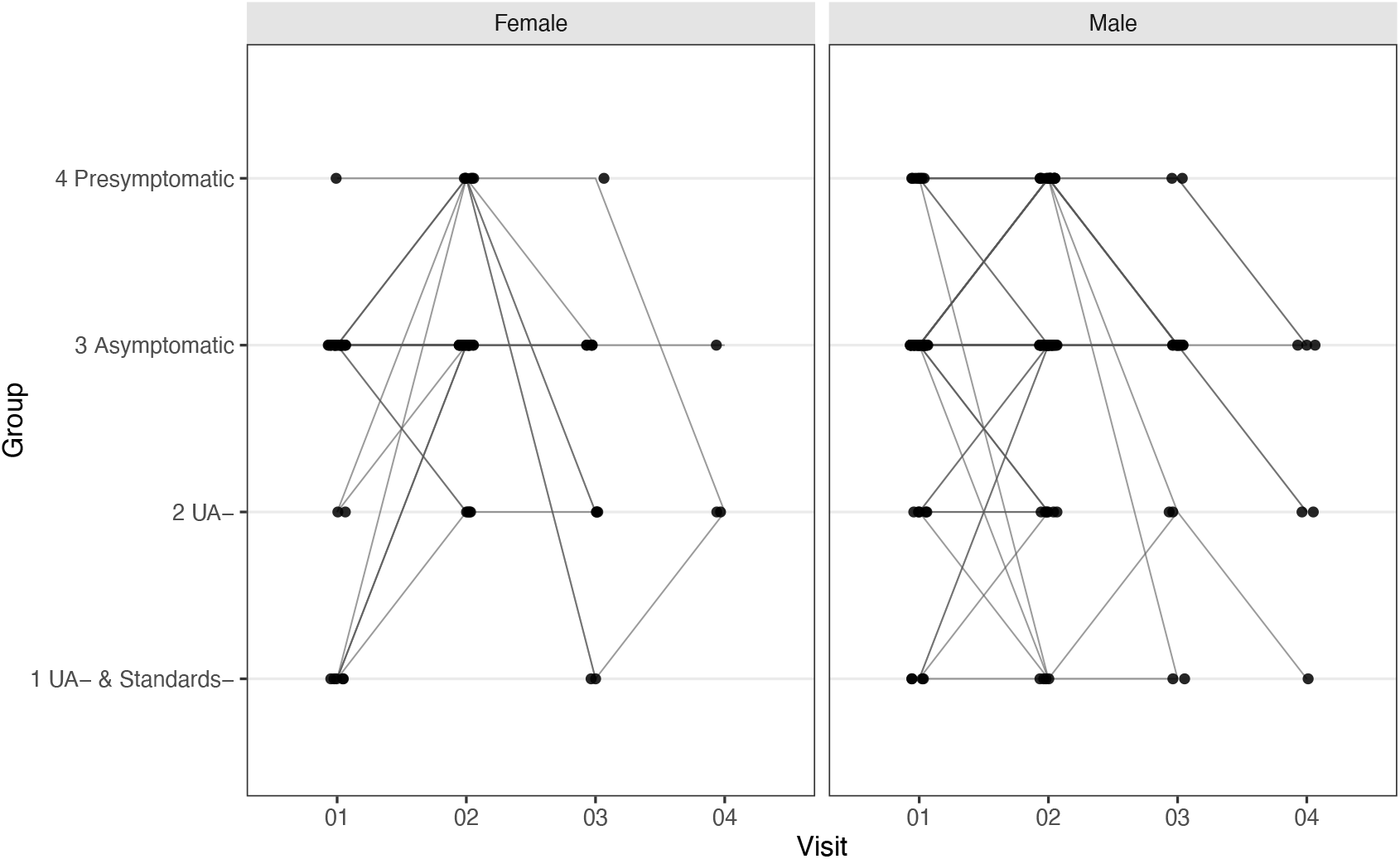
Within-person variation in urine sample characteristics (Presymptomatic (Level 4), Asymptomatic (Level 3), UA-Negative (“UA-”, Level 2), or UA and Standards Negative (“UA-& Standards -”, Level 1) over time by sex. Consecutive Asymptomatic not shown; the four groups as shown are mutually exclusive and exhaustive.

Considering Figure 2, 10/33 (30.3%) sample pairs from females and 15/61 (24.6%) sample pairs from males met different Group definitions over time when all samples were 72 hours asymptomatic. Being presymptomatic at one visit did not result in a higher likelihood of positive nitrites, LE, WBC, or positive SUC results at a subsequent visit (2 weeks later). However, the majority of observations in Figure 2 show that, once an individual had a presymptomatic sample, the next sample was most often classified in the same group as the sample prior to being presymptomatic.

## Discussion

Our study demonstrates that in the absence of symptoms, the majority of people with NLUTD due to SCID and who manage their bladders with IC have inflammation (6+WBC/hpf), bacterial growth on culture consistent with historical identification of UTI, and uNGAL values greater than those of the healthy non-NLUTD population. The wide variability of these standard urinary markers in various asymptomatic states limits their utility in clinical decision making around UTI.

In addition, we observed substantial between-person variability in standard urine markers — including LE, nitrite, urine WBC count, and bacteriuria —among samples from individuals who were asymptomatic for 72 hours after the sample (within-person variability not shown). This variability further undermines the assumption that these markers can reliably characterize urinary health in individuals with NLUTD when measured at one timepoint and with no other information, including a full assessment of validated symptoms. Therefore, consideration should be given to individualized interpretation of urinary markers in the context of baseline variability and sex, rather than utilizing clinical thresholds that are assumed to apply universally or be meaningfully “statistically controlled” for the sex of the individual.

A small proportion of the participants met the criteria for UA negative+SUC negative urine (Standards negative) or UA negative only. This reiterates that baseline inflammation and bacterial growth is present in the vast majority of people with NLUTD due to SCI/D and who manage their bladders with IC irrespective of symptoms. Rather than one-time urinary markers, a change from baseline and possibly different urinary biomarkers to inform UTI should be considered. Our results suggest that since urinary markers vary so widely among individuals who are asymptomatic at the time of the urine sample, individuals with symptoms at the time of assessment are likely to have at minimum similar proportions of positive inflammation, bacterial growth and uNGAL levels. These findings challenge the clinical utility of routine UA and culture-based markers at a single time point, regardless of symptom status, for diagnosing or monitoring urinary health in individuals with SCI/D. Reliance on these markers may lead to misclassification, overtreatment, or both in this population.

These results support several best practice statements. Prior to urologic procedures that breach the urinary mucosa, in accordance with the American Urological Association (AUA)’s Best Practice Statement on Urologic Procedures and Antimicrobial Prophylaxis^21^ and the 2019 Infectious Disease Society of America asymptomatic bacteriuria guideline^22^, patients with NLUTD should undergo a pre-procedure urine culture and receive a short-course (typically single-dose), culture-directed antimicrobial prophylaxis approximately 30-60 minutes pre-procedure (and not a multi-day “therapeutic” regimen). In these statements, it is emphasized that procedure-associated infection risk is driven by the procedure, not by the host. For urodynamic studies, the best practice policy from The Society of Urodynamics, Female Pelvic Medicine and Urogenital Reconstruction recommends that high-risk patients be screened for symptomatic infection, and when prophylaxis is used, received only a single oral dose of trimethoprim-sulfamethoxazole (or alternative if allergic) before testing^23^. One study has assessed single dose vs longer course (3-5 days) of pre-procedural antibiotics prior to endoscopic urological procedures in people with SCI/D and asymptomatic bacteriuria.

Among 60 participants, there were no significant differences in vital signs, leukocytosis, adverse events and satisfaction among participants in the two groups. Cost and pre-procedure anxiety was greater for individuals in the 3-5 day antibiotic group^24^. Our results demonstrating that bacterial growth in the absence of symptoms (asymptomatic bacteriuria) occurs in 58.4%-63.3% of cases, and that even in those with absent symptoms and negative urinalysis, 12.0-36.8% might have significant growth on SUC, support these best practice statements.

We also noted significant variability in uNGAL values among asymptomatic individuals and across sex. The most recent reference ranges for uNGAL, reported for healthy adults without (SCI/D)^25^ (2012) have a 95th percentile value of 107 µg/L. Tables 4-5 show that the reference range for uNGAL for individuals with NLUTD due to SCI/D far exceeds this 2012 95th percentile value. Also, Table 4 shows that the reference range values depend critically on both the definition of “asymptomatic” used, and also the sex of the individual However, the range of uNGAL in samples in the Pre-symptomatic group from males (9.1 - 996.4) was much higher than that of the Standards-Negative group from males (4.9-74.2), while the range of uNGAL for the Pre-symptomatic group from females (5.1-166.0) was much smaller than that of their Standards-Negative samples (3.1 - 484.8). This observed range is considerably wider than has been reported for other adult populations^25^.

In this work, we did not normalize uNGAL by urine creatinine, but this is unlikely to have caused the very wide range and high variability in uNGAL values across samples.

Similar to uNGAL, we also observed a wide range in the distribution of uWBC across individuals in each of the classifications of “asymptomatic” (Table 2). Again, the proportions of participants whose samples exhibited 0-5 WBC by urinalysis varied by sex and group. Comparatively, we saw almost no differences between the proportions of samples from men and women that were negative for nitrites, except for the two definitions that specified UA nitrite levels to be negative (i.e., all were 100% negative by definition); 40-50% of all samples from individuals who were asymptomatic for 72 hours before and after the time of the urine sample had positive nitrites. Finally, we also found a large proportion of urine samples from men and women who were asymptomatic for 72 hours after the sample on at least one, and on two consecutive, occasions yielded at least one organism at 10^5^ CFU. These findings highlight the variability within these markers in asymptomatic people with SCI/D.

This study is limited in that we did not include any cases where a UTI developed; moreover, “presymptomatic” samples were assigned that classification when even one USQNB symptom was endorsed within 72 hours of the sample. Other limitations include lack of assessment of renal status, which is a potential confounder of uNGAL. Despite these limitations, our study had several strengths, most notable the repeated samples, our strict definitions of “asymptomatic”, and stratification of asymptomaticity based on durability of absence of symptoms and laboratory findings.

## Conclusions

This study of individuals with NLUTD due to SCI/D and who manage their bladders with IC demonstrates substantial variability in urinary markers in people who were asymptomatic for 72 hours before and after sampling. Given the wide range of urine WBC counts, nitrite results, and bacterial growth observed even in the absence of bladder, urine, and constitutional symptoms, it is unlikely that the presence of these findings at any level alone meaningfully informs clinical decision making. Although the combined absence of both inflammation and bacterial growth may help to exclude UTI diagnosis, this pattern occurs in only a small minority of cases. These observations suggest that alternative approaches to UTI diagnosis in this population may be warranted, such as symptom-driven algorithms, individualized change-from-baseline criteria, or the incorporation of novel biomarkers.

## Data Availability

All data produced in the present study are available upon reasonable request to the authors

## References Cited

1. Crescenze IM, Lenherr SM, Myers JB, et al. Self-Reported Urological Hospitalizations or Emergency Room Visits in a Contemporary Spinal Cord Injury Cohort. J Urol. 2021;205(2):477–482. doi:10.1097/JU.0000000000001386

2. Stampas A, Dominick E, Zhu L. Evaluation of functional outcomes in traumatic spinal cord injury with rehabilitation-acquired urinary tract infections: A retrospective study. J Spinal Cord Med. 2019;42(5):579–585. doi:10.1080/10790268.2018.1452389

3. Skelton F, Salemi JL, Akpati L, et al. Genitourinary Complications Are a Leading and Expensive Cause of Emergency Department and Inpatient Encounters for Persons With Spinal Cord Injury. Arch Phys Med Rehabil. 2019;100(9):1614–1621. doi:10.1016/j.apmr.2019.02.013

4. Skelton-Dudley F, Doan J, Suda K, Holmes SA, Evans C, Trautner B. Spinal Cord Injury Creates Unique Challenges in Diagnosis and Management of Catheter-Associated Urinary Tract Infection. Top Spinal Cord Inj Rehabil. 2019;25(4):331–339. doi:10.1310/sci2504-331

5. Forster CS, Miller RG, Gibeau A, et al. Accuracy of Urinalysis for UTI in Spina Bifida. Pediatrics. 2024;154(1):e2023065192. doi:10.1542/peds.2023-065192

6. Tractenberg R, Aguirre Guemez A, Petriello M, Groah S. SCI Model Systems Complicated UTI (cUTI) Guidelines: International consensus on likelihood of symptomatic cUTI. Top Spinal Cord Inj Res. Published online In Press 2025.

7. The Prevention and Management of Urinary Tract Infections Among People With Spinal Cord Injuries. J Am Paraplegia Soc. 1992;15(3):194–207. doi:10.1080/01952307.1992.11735873

8. Hooton TM, Bradley SF, Cardenas DD, et al. Diagnosis, Prevention, and Treatment of Catheter-Associated Urinary Tract Infection in Adults: 2009 International Clinical Practice Guidelines from the Infectious Diseases Society of America. Clin Infect Dis. 2010;50(5):625–663. doi:10.1086/650482

9. Bonkat G, Wagenlehner F, Kranz J. Keep it Simple: A Proposal for a New Definition of Uncomplicated and Complicated Urinary Tract Infections from the EAU Urological Infections Guidelines Panel. Eur Urol. 2024;86(3):195–197. doi:10.1016/j.eururo.2024.05.007

10. Craven BC, Alavinia SM, Gajewski JB, et al. Conception and development of Urinary Tract Infection indicators to advance the quality of spinal cord injury rehabilitation: SCI-High Project. J Spinal Cord Med. 2019;42(Sup1):205–214. doi:10.1080/10790268.2019.1647928

11. Madden-Fuentes RJ, McNamara ER, Lloyd JC, et al. Variation in definitions of urinary tract infections in spina bifida patients: a systematic review. Pediatrics. 2013;132(1):132–139. doi:10.1542/peds.2013-0557

12. DiSabato DJ, Marion CM, Mifflin KA, et al. System failure: Systemic inflammation following spinal cord injury. Eur J Immunol. 2024;54(1):2250274. doi:10.1002/eji.202250274

13. Rounds AK, Tractenberg RE, Groah SL, et al. Urinary Symptoms Are Unrelated to Leukocyte Esterase and Nitrite Among Indwelling Catheter Users. Top Spinal Cord Inj Rehabil. 2023;29(1):82–93. doi:10.46292/sci22-00095

14. Groah SL, Pérez-Losada M, Caldovic L, et al. Redefining Healthy Urine: A Cross-Sectional Exploratory Metagenomic Study of People With and Without Bladder Dysfunction. J Urol. 2016;196(2):579–587. doi:10.1016/j.juro.2016.01.088

15. Fitzpatrick MA, Wirth M, Burns SP, et al. Management of Asymptomatic Bacteriuria and Urinary Tract Infections in Patients With Neurogenic Bladder and Factors Associated With Inappropriate Diagnosis and Treatment. Arch Phys Med Rehabil. 2024;105(1):112–119. doi:10.1016/j.apmr.2023.09.023

16. Wickham A, McElroy SF, Austenfeld L, et al. Antibiotic use for asymptomatic bacteriuria in children with neurogenic bladder. Brei T, Castillo H, Castillo J, Thibadeau J, eds. J Pediatr Rehabil Med. 2022;15(4):633–638. doi:10.3233/PRM-210051

17. Tractenberg RE, Groah SL, Rounds AK, Ljungberg IH, Schladen MM. Preliminary validation of a Urinary Symptom Questionnaire for individuals with Neuropathic Bladder using Intermittent Catheterization (USQNB-IC): A patient-centered patient reported outcome. PLOS ONE. 2018;13(7):e0197568. doi:10.1371/journal.pone.0197568

18. Jones-Freeman B, Chonwerawong M, Marcelino VR, Deshpande AV, Forster SC, Starkey MR. The microbiome and host mucosal interactions in urinary tract diseases. Mucosal Immunol. 2021;14(4):779–792. doi:10.1038/s41385-020-00372-5

19. IBM Statistics for Windows. Published online 2013.

20. Posit team. RStudio: Integrated Development Environment for R. Published online 2025. http://www.posit.co/

21. Lightner DJ, Wymer K, Sanchez J, Kavoussi L. Best Practice Statement on Urologic Procedures and Antimicrobial Prophylaxis. J Urol. 2020;203(2):351–356. doi:10.1097/JU.0000000000000509

22. Nicolle LE, Gupta K, Bradley SF, et al. Clinical Practice Guideline for the Management of Asymptomatic Bacteriuria: 2019 Update by the Infectious Diseases Society of America. Clin Infect Dis. 2019;68(10):e83–e110. doi:10.1093/cid/ciy1121

23. Cameron AP, Campeau L, Brucker BM, et al. Best practice policy statement on urodynamic antibiotic prophylaxis in the non-index patient. Neurourol Urodyn. 2017;36(4):915–926. doi:10.1002/nau.23253

24. Chong JT, Klausner AP, Petrossian A, et al. Pre-procedural antibiotics for endoscopic urological procedures: Initial experience in individuals with spinal cord injury and asymptomatic bacteriuria. J Spinal Cord Med. 2015;38(2):187–192. doi:10.1179/2045772313Y.0000000185

25. Cullen MR, Murray PT, Fitzgibbon MC. Establishment of a reference interval for urinary neutrophil gelatinase-associated lipocalin. Ann Clin Biochem Int J Lab Med. 2012;49(2):190–193. doi:10.1258/acb.2011.011105

